# Reduction in transfer of micro-organisms between patients and staff using short-sleeved gowns and hand/arm hygiene in Intensive Care during the Covid pandemic: a simulation-based randomised trial

**DOI:** 10.1101/2021.01.16.21249221

**Authors:** Laura Vincent, Mudathir Ibrahim, Joanne Kitchin, Claire Pickering, Jennie Wilson, Enrico Sorrentino, Claudia Salvagno, Laurie Earl, Louise Ma, Kathryn Simpson, Rose Baker, Peter McCulloch

## Abstract

**Background:** Current PPE practices in UK intensive care units involve “sessional” gown use. This protects staff, but puts patients at risk of nosocomial infection via PPE gowns. Anecdotal reports of such infections in ITUs during Covid are frequent. We therefore explored the use of short-sleeved gowns with hand and arm hygiene as an alternative to sessional gowns.

**Methods:** ITU Staff were invited for simulation suite training in Covid intubation and proning. They were trained in a specific hand and arm washing technique before performing simulated tasks using both standard and modified (short sleeved) PPE. Fluorescent powder was used to simulate micro-organisms, and detected using standardised photos under U/V light. Teams of staff were randomised to use standard or modified PPE first. Individuals were questioned about their feeling of personal safety, comfort and the patient’s safety at 4 intervals.

**Results:** 68 staff and 17 proning volunteers were studied in 17 sessions. Modified PPE completely prevented staff contamination during Covid intubation, which occurred in 30/67 staff wearing standard PPE (p = 0.0029, McNemar). Conversely, proning volunteers were contaminated by staff in 15/17 sessions with standard PPE and in 1/17 with modified PPE (P = 0.0227 McNemar). Impressions of staff comfort were superior with modified PPE (p< 0.001, t-test); personal safety scored higher with standard PPE, but the difference decreased during the session (p<0.001 start, 0.068 end). Impressions of patient safety were initially similar (p=0.87) but finished strongly in favour of modified PPE (p<0.001).

**Conclusions:** Modified PPE using short sleeves and hand/arm cleansing appears superior to standard PPE with sessional gowns in preventing transfer of contamination between staff and patients. A clinical trial of this strategy is merited.

## BACKGROUND

Since the early stage of the COVID19 pandemic, a priority for healthcare professionals across the world has been the safety, availability and accessibility of Personal Protective Equipment. As our understanding of the epidemiology and pathophysiology of COVID19 has grown, national guidance from infection control experts, including Public Health England, has evolved in response. Now known to be a virus transmitted via the respiratory tract, the level of protection required has been reduced and is distinct from that of a High Consequence Infectious Disease such as Ebola. By virtue of both perceived and genuine inequity in PPE provision and the understandable fear of healthcare professionals of contracting the virus through occupational exposure, PPE has been a focus of significant anxiety and psychological stress throughout the pandemic. This has particularly been the case when changes in guidance have recommended a ‘downgrading’ of the PPE.

Recommended precautions are based on the potential route of transmission^1,2^. When healthcare professionals are caring for patients undergoing ‘Aerosol Generating Procedures’ (procedures that result in the production of airborne particles, which can remain suspended in the air, travel over a distance and cause infection if inhaled) a higher level of protection is required, ‘Level 2 PPE’^3^. Whilst facial protection is fairly consistent country wide, the types of gown worn and the number and combination of gloves varies considerably. Guidance on this is not definitive and there is scanty evidence to date for ‘best practice’.

During wave 1 of the COVID19 pandemic, there were widespread reports from ICUs across the UK of rising rates of healthcare acquired infections with multi-resistant organisms (personal communication, P Dean, PPE lead for National Emergency Committee for Critical Care). It was inferred by infection control experts that the sessional use of long sleeved gowns, worn by staff looking after multiple high dependency patients in cohorted areas was likely a significant contributory factor. In addition, due to adherence to PPE protocols, staff were not washing their hands between patient care episodes, which would usually be standard practice.

A team of infection control, human factors, intensive care and education experts has collaborated to develop a new protocol for Level 2 PPE, aiming to reduce the risk of healthcare staff mediated transmission of multi-resistant organisms.

It was acknowledged that the safety of staff is paramount, so any change in PPE must be demonstrated to be equally as effective at protecting staff from infection as that which they are currently using and importantly, no changes would be made to facial protection.

The importance of comfort and psychological safety of staff looking after patients with COVID 19 was also recognised, particularly when PPE is worn for an extended period of time. Any changes must therefore be made adopting a human factors approach, taking into consideration perceived as well as actual changes in degree of protection and hence feelings of vulnerability. For any change to be effective, engagement of staff in the understanding and the process of change is as important as the data obtained from the study. A simulation based experiment was therefore designed, allowing us to compare the effectiveness of the two Level 2 PPE protocols (control and experimental) at protecting staff from ‘residual contamination’ after doffing and protecting patients from contamination through staff and patient contact.

## METHODS

### Objectives

The primary objective of this study was to develop and test new protocols for Level 2 PPE, which are specifically designed to reduce the spread of multi-drug resistant organisms in ICU environments, whilst maintaining a level of protection from COVID19 equivalent to current protocols.

### Outcome measures

The primary outcome of this study was contamination of staff and simulated patients with fluorescent powder when staff wore adapted (experimental) Level 2 PPE in comparison to the current (control) Level 2 PPE.

The secondary outcomes were:

∘ Staff perceptions of comfort, personal safety and safety of the patient when wearing experimental versus control Level 2 PPE, during simulation tasks
∘ Changes in staff perceptions of comfort and safety wearing each type of PPE when questioned (a) before the PPE training, (b) after the PPE training and (c) after the simulation training exercises.

### Study Design

We conducted a dual-centre, non-blinded randomised cross-over trial in Oxford University Hospitals (OUH), Oxford and Whittington Hospital, London. This was preceded by an uncontrolled cohort study of the development of the new PPE protocol, based on the recommendations for IDEAL Stage 2a and 2b studies

Ethical approval for this trial was obtained from Oxford University Research Ethics Committee (CUREC) (Reference: R72882/RE001).

Participants were nurses, physiotherapists or doctors working in Adult Intensive Care Units at Oxford University Hospitals Trust and the Whittington Health NHS Trust (London). Participants were recruited by an email invitation from the investigators. Prior to enrolment, written informed consent was obtained, which included specific reference to the use of photography and video recording.

#### Development of Level 2 PPE protocol

Guided by infection prevention and control experts, the study team determined the principle changes to be made, namely, transition from a long sleeve to a short sleeve gown and from double gloves to single gloves. No changes were made to facial protection.

In the new protocol, after any patient interaction, staff should remove their plastic apron and single pair of gloves, wash their hands and forearms and don a new apron and gloves prior to attending another patient. This process was termed a ‘mini-doff’, to differentiate from the ‘full doff’ undertaken when leaving a Level 2 COVID cohorted area.

Prototype Level 2 PPE doffing and mini-doffing protocols were designed and revised through an iterative co-design process following the recommendations for IDEAL Stage 2a studies^4^. Members of ICU staff were invited to test the prototype protocols, feeding back to the study team on their subjective comfort and safety and the ease of use of the associated visual aids, until a stable version was established.

Use of ultraviolet tracer demonstrated that the proposed protocols were effective, but precision in application, including a meticulous hand and forearm washing process, was necessary to ensure complete decontamination. In view of this, and to avoid bias in the study due to learning curve effects with the new PPE, all subjects for the randomised study underwent preliminary training. This involved demonstration and repeated practice of current and experimental Level 2 PPE and the respective doffing and mini-doffing protocols until competence was demonstrated in both, following the principles of IDEAL Stage 2b studies^4^.

#### Level 2 PPE content and protocols

##### Level 2 PPE Control

The ‘Control’ Level 2 PPE protocol is that which is currently employed in the respective hospitals. In OUH this consisted of FFP3 mask, hat, visor, long sleeved gown, plastic apron, long sleeved surgical gloves (inner) and short sleeve gloves (outer) In the Whittington hospital the long sleeves gloves were not used (single gloving). In both hospitals standard practice was to change the apron and one pair of gloves between patients.

##### Level 2 PPE Experimental

The ‘Experimental’ Level 2 PPE protocol consisted of FFP3 mask, hat, visor, short sleeved gown, plastic apron, short sleeve gloves (single pair). The new “mini-doffing” protocol for moving between patients included removal of apron and gloves, thorough washing of hands and arms and donning of a fresh apron and gloves. The handwashing technique taught emphasised the need to rinse and dry the arms with the hand hanging down, and all washing and drying movements directed from the elbow distally, as the IDEAL 2a study showed that this avoided contamination around the elbows.

#### Randomised Study

After training, participants were divided into groups of 4 (5 in the Whittington), each containing at least one airway-trained doctor. Each group performed 2 simulated activities (Covid intubation and proning) twice – once with control PPE and once with the experimental PPE. The order in which the protocols were used was randomised in permuted blocks of four sessions, using a computer generated random number These simulated training tasks were selected to ensure significant contact with the ‘patient’ and to prepare staff entering the second wave of the pandemic. They were conducted and debriefed as a normal training session. The study teams in OUH and The Whittington hospital used their local protocols for each activity to maximise the training impact.

Simulation task 1 was oral endo-tracheal intubation of a simulated patient (a high-fidelity mannequin), with hypoxaemic respiratory failure, secondary to suspected COVID19 pneumonia. Participants were expected to proceed with intubation using the bespoke checklists of their respective hospital. They were also asked to change the patient’s bed sheet and hospital gown immediately following intubation to increase the amount of contact with the patient.

Simulation task 2 was turning a simulated patient (an actor) from the supine position (lying on their back) into the prone position (lying on their front), which is a standard therapy for ventilated patients with refractory hypoxaemic respiratory failure. Participants were expected to proceed using the bespoke proning checklist of their respective hospital. Prior to proning, they were also asked to change the patients undersheet to increase contact with the patient.

Since the proning guideline requires a fifth team member, a volunteer joined the team to facilitate completion of this task but was not included in any data collection.

An ultraviolet tracer powder (Glogerm, www.Hygienicsolutions.com), which glows white under ultraviolet light, was used to study the effectiveness of the control and experimental Level 2 PPE. The powder was used to represent contamination of staff and patients with both COVID 19 and organisms which may be spread by skin to skin contact.

Bespoke plastic scoops (internal volume approximately 0.4 cm^3)^ were made to ensure application of a consistent mass of powder, which was applied and distributed by the gloved hands of the study team.

Prior to simulation task 1, powder was applied to the simulated patient (mannequin) − 2 scoops to the head, 3 scoops to the ventral aspect of the arms, 2 scoops to the chest, 2 scoops to the mannequin’s gown and 2 scoops to the blanket.

Prior to simulation task 2, powder was applied to the arms of the participants – 3 scoops to each arm (including hands and both ventral and dorsal aspects).

#### Power calculation

Sample size calculations were based on data analysis using the mid-P version of the exact McNemar’s test. Power was calculated by finding the percentage of 100,000 simulated datasets for which the null hypothesis would be rejected at the 5% level of significance with β = 0.9. This indicated a minimum of 50 participants would be required.

### Conduct of simulation sessions

At the start of the simulation sessions the team received a briefing on the session and simulation tasks, and an introduction to the mannequin. The sessions then proceeded according to the flowchart (see Figure 1).

**Figure 1.**
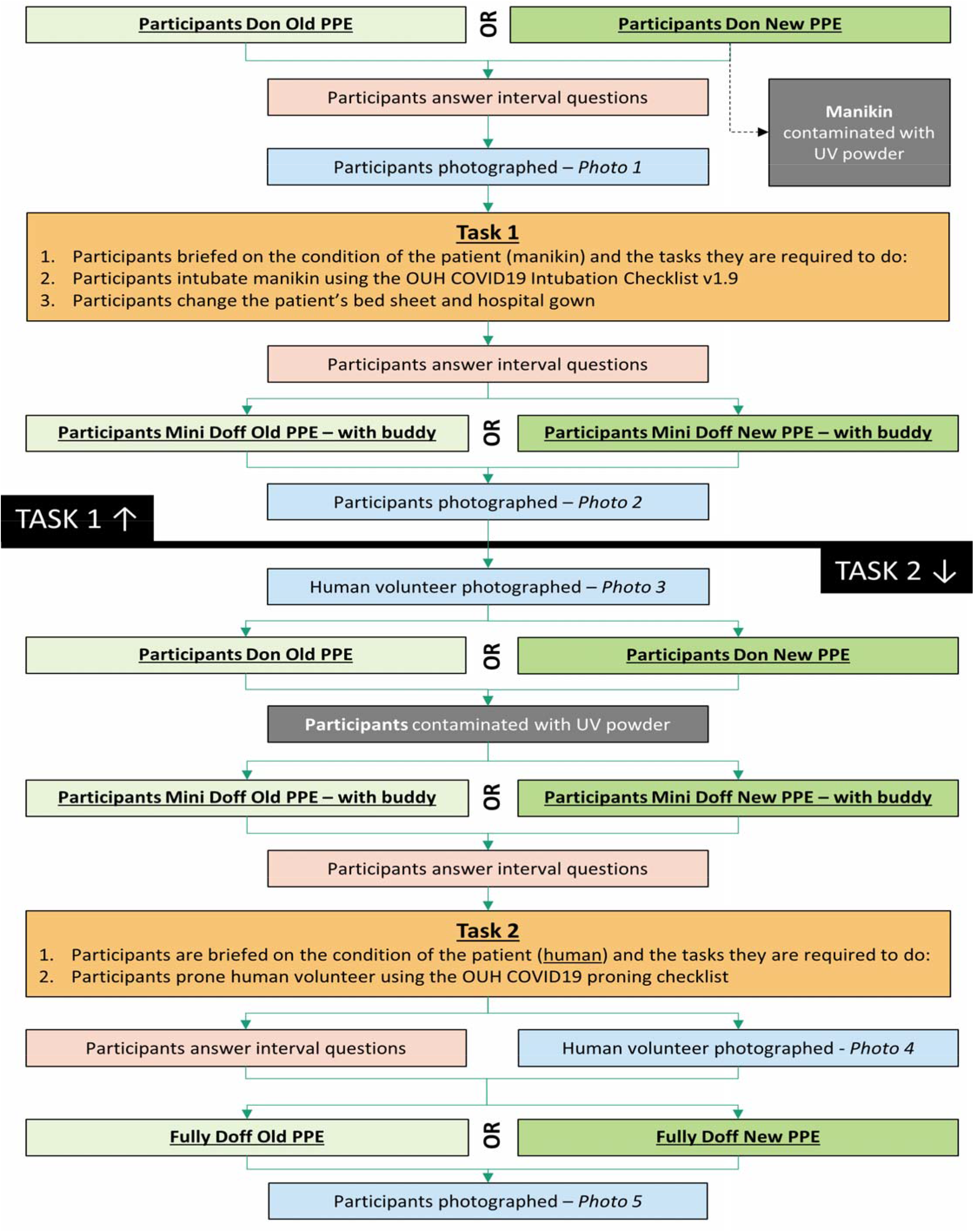
Process map for Simulation sessions.

Participants first donned the Control or Experimental Level 2 PPE according to randomisation. A photograph was taken under UV light to assess for any baseline contamination (expected to be zero). They then undertook Simulation Task 1 (intubation), following which they performed a ‘mini-doff’ and were photographed again, to assess the extent of contamination on their hands (or gloves) and forearms (or sleeves).

Having replaced their apron and gloves, powder was then applied to the participants hands and forearms and they were asked to ‘mini-doff’ again, as if moving between patients. After replacing their apron and gloves, they then undertook Simulation task 2 (proning), after which they were asked to undertake a ‘full doff’. A 3rd photograph was taken at this point to assess for any residual contamination. The ‘proned’ volunteer wore fresh scrubs for every proning episode and was photographed before and after being proned to assess for contamination.

A simulation debrief was performed after task 2, and the whole process was then repeated wearing the other version of Level 2 PPE.

### Data collection

#### Questionnaires

Simple questionnaires were used at 4 intervals before, during and after the simulation session to evaluate participants’ feelings about their personal safety, physical comfort and the safety of their “patient” on a 10 point Likert scale. (See Appendix 1)

#### Photography

Participants were photographed twice at each photography interval, first facing forward with arms up and palms facing forward and second facing forward with palms facing backwards. The ‘proned’ patient was photographed before and after each proning exercise, from in front and from behind.

The camera, light and subject positions were standardised to ensure consistent data was obtained. The camera was positioned to allow the arms and torso to be captured. The UV lamp was placed close to camera height and as far from the participant as possible. Positioning markers for feet and index fingers ensured participants stood in exactly the same place and position in front of a black screen with a black and white chequerboard attached to facilitate analysis.

### Data Analysis and Statistics

Photographs were assessed by two independent observers. For participant photos, attention was focused on the forearm area below the elbow, where the two protocols differed. For the proning volunteer the whole body was studied from anterior and posterior views. Photos were assessed as either showing contamination or none. Digital quantitation of the area of contamination was performed and will be reported in further communications. Differences in the percentage of participants showing contamination were evaluated using McNemar’s test to compare Baseline (pre-activity) Post-intubation and Post-full doffing contamination for the Experimental and Control PPE protocols in a modified intention to treat analysis.

Staff impressions of comfort, personal safety and patient safety were compared using single sample t-tests to compare mean differences between the Experimental and Control groups at each time interval. Changes in staff views between the beginning and end of the study were evaluated using both Pearson and Spearman (ranked) correlation between score and task number. All tests were 2 sided.

## RESULTS

### Ideal Studies

The IDEAL stage 2a study was carried out using research team members and staff volunteers as participants. Ideas to improve the protocol were tested in 4 sessions which resulted in 5 iterative changes:

1. Washing all the way to the elbows was emphasised, as initially it was not properly carried out by some participants
2. A surgical scrubbing up technique was adopted, but then abandoned, as it left a ring of contamination just above the elbows.
3. The sleeves of the short sleeved gown were rolled up to ensure they did not get wet or interfere with washing
4. A technique involving rinsing and drying from the elbows distally was adopted to deal with the “elbow ring” problem, and proved effective in achieving 100% removal of contamination when properly carried out.
5. Several iterations of the visual aids were produced until a satisfactory version was achieved.

The IDEAL Stage 2b study was conducted informally, assessing the performance of all participants during training in doffing and donning. Errors in washing technique were initially frequent but decreased rapidly with practice and coaching, and were virtually eliminated in most participants after 3 attempts. No formal analysis of learning curves was attempted.

### Randomised Trial

Thirteen simulation sessions were held at OUH and 4 at the Whittington. Four participants dropped out of sessions at OUH and had to be substituted by research team members, leaving 48 participants at OUH and 20 at the Whittington (who used a team of 5 people per session).

#### Contamination Results

68 participants and 17 proning volunteers were studied. One participant in the Control group was excluded as no photos could be found except the baseline picture, leaving 67 participants for analysis (see Table 1). 2 participants had detectable contamination at baseline wearing the Experimental PPE, and none while wearing the Control PPE. After the Intubation scenario 30 of 67 participants had detectable contamination wearing the Control PPE (on the sleeves of the retained gown) and none wearing the Experimental PPE (p < 0.0001, McNemar). After proning by participants wearing Control PPE, contamination of the ‘patient’ was detected in 15 of 17 episodes, compared to 2 of 17 episodes for the Experimental PPE (P < 0.001, McNemar). At final doffing, 18 of 67 participants in the Experimental group showed contamination compared to 7 of 67 in the Control group (p = 0.0116).

**TABLE 1.** Presence of contamination as shown by UV photography. Numbers represent participants and proning volunteers. Participants n = 67, Volunteers n = 17.

|  | Participants<br>EXPERIMENTAL |  | Participants<br>CONTROL |  | Proning Volunteer<br>POST PRONING |  |
| --- | --- | --- | --- | --- | --- | --- |
|  | Post<br>Intubation | Post<br>Proning | Post<br>Intubation | Post<br>Proning | EXP. | CONTROL |
| Y | 0 | 18 | 30 | 7 | 1 | 15 |
| N | 67 | 49 | 37 | 60 | 16 | 2 |

#### Questionnaire Data

Feelings of personal comfort were consistently better in the Experimental PPE than in the Control PPE (See Table 2). Personal safety was scored lower with the Experimental PPE, but the difference between the groups decreased as participants gained experience of the Experimental equipment (p = 0.002, Spearman). The participants’ impression of the safety of the patient also changed in favour of the Experimental PPE during the simulation, and it was strongly preferred from this point of view by the end of the session (p< 0.001, Spearman).

**Table 2.** Participating Staff impressions of personal comfort, personal safety and patient protection against infection during Simulation. Values are mean differences between Likert scale scores, 95% confidence intervals and t-test results.

|  | Before Intubation | After Intubation | Before Proning | After Proning |
| --- | --- | --- | --- | --- |
| Comfort | 2.125<br>(1.539-2.711)<br>t = 7.3, p < 0.001 | 2.67<br>(2.004 – 3.329)<br>t = 8.1, p < 0.001 | 2.063<br>(1.34 – 2.785)<br>t = 5.74 p < 0.001 | 2.75<br>(1.954 – 3.546)<br>t = 6.95, p < 0.001 |
| Personal Safety | -1.646<br>(-2.116 to -1.176)<br>t = -7.05, p < 0.001 | -0.958<br>(-1.406 to -0.51)<br>t = -0.43, p < 0.001 | -0.604<br>(-1.159 to -0.05)<br>t = -2.19, p = 0.033 | -0.625<br>(-1.299 to 0.05)<br>t = -1.87, p = 0.068 |
| Patient Protection | -0.042<br>(-0.55 to +0.466)<br>t = -0.196, p = 0.87 | 1.104<br>(0.472 – 1.736)<br>t = 3.52, p = 0.001 | 2.042<br>(1.458 – 2.625)<br>t = 7.04, p < 0.001 | 2.854<br>(2.193 – 3.516)<br>t = 8.68, p < 0.001 |

## DISCUSSION

The purpose of this study was to determine whether a PPE protocol using short sleeved gowns and hand/arm hygiene could protect staff from contamination whilst reducing the risk of spreading nosocomial infections. A simulation study was performed because a study in a live clinical setting was not feasible due to staff concerns, ethical concerns and statistical power challenges. An initial development and learning phase based on the IDEAL Framework was used to avoid bias or disruption of the randomised study due to learning curve effects or changes in the Experimental protocol. Randomisation of the order of use of the PPE protocols was performed because of an obvious learning effect amongst teams, which were invariably faster and made less errors on their second attempt. This is well recognised in evaluation of simulation work^5^. The simulation study was designed in accordance with the well-established template for ICU multi-disciplinary simulation training in the OxSTaR simulation centre. The learning objectives fulfilled the current clinical educational priorities for ICU staff entering the 2^nd^ wave of the COVID19 pandemic. The format was consistent with our usual practice, comprising a safety briefing, a standardised high-fidelity simulation scenario and a structured debrief, led by an experienced trained facilitator.

The results demonstrated a very large reduction in the probability of acquired contamination on the arm and hand areas for the Experimental PPE protocol after the ‘mini-doff’ and hand/forearm hygiene routine between patients had been performed, compared with the control PPE protocols in which the long-sleeved gown is not changed. The proning simulation confirmed that such contamination was readily transferred to other patients during procedures requiring manual handling of the patient, and was greatly reduced (from 88% to 6% of simulated pronings) using the Experimental PPE protocol. The handwashing method was very effective but required significant training and careful attention. This probably explains the one discordant result, that staff wearing the Experimental PPE had more contamination evident following the full doff after proning than with Control PPE. Since the volunteer was not contaminated at the start of the proning simulation, the contamination found can only have come from the participants’ own arms, which we deliberately loaded with a high dose of powder to ensure we could detect transfer to the volunteer. Proning requires staff to push the upper forearms under the patient, which would tend to push excess powder up towards the elbow, and improper use of the washing protocol will result in a ring of contamination at the elbow in these circumstances, as our IDEAL 2a study had shown. We reviewed the photographs of the 18 Experimental cases of participant contamination post-proning and found this pattern in all 18, with only very slight contamination elsewhere. It is also notable that only 1 volunteer was contaminated in this part of the study.

Human Factors principles suggest that reliance on precise attention to detail in routine but complex tasks is likely to lead to significant failure rates^6,7^. Although very satisfactory as a simulation exercise, the washing protocol used here as a demonstration might be impractical in clinical practice in a busy ICU. There is a large body of evidence that alcohol gel application is equally effective^8,8,10^, and a clinical version of the protocol would likely substitute this for the hand washing techniques, eliminating the problem.

Our study suggests that short sleeved gowns with the use of hand and arm hygiene between patients is as effective as sessional long-sleeved gowns at avoiding staff contamination, and could considerably reduce the risk of transmission of nosocomial infection via gown sleeves. We therefore believe that a clinical trial of the new protocol is merited. Staff comfort was significantly better in the Experimental PPE, and their confidence in both their own safety and that of the patient improved steadily during their participation in the study. This suggests that reluctance of staff to adopt the new PPE might be overcome by introducing it via simulation training.

This simulation study could not reliably reproduce the behaviour of either the Covid 19 virus or bacteria, and it is possible that these might be more difficult (or easier) to eradicate. The analysis of photographs could not be blinded, since the observer could see which type of gown participants wore, but the binary yes/no choice between any contamination and none by independent observers was effectively objective. Given the urgency of the current situation, we suggest that introduction of the new PPE using a stepped wedge design trial to stage its’ introduction in ICUs or Trusts might allow both high quality evaluation of the new PPE and facilitated rapid adoption.

## Data Availability

Once the article has been accepted by a peer reviewed journal, all relevant data will be deposited in the Oxford University Research Archive (ORA) at https://ora.ox.ac.uk/

## ACKNOWLEDGEMENTS

We would like to thank Dr Katie Jeffery and Lisa Butcher (Microbiology & Infection Control, OUHFT) for helpful advice and Dr David Garry, Lyn Bennet and Mr Trevor Venes (Clinical Lead, Matron and Nursing Education lead for ITU, OUHFT) for their strong support and help with recruitment. Thanks also to the staff of the OxStar simulation suite (OUHFT). At the Whittington Hospital we thank Anezka Pratley (CRN), Ana Alvaro, Karen Johnston and the staff of the Simulation Centre.

Thanks also to Professor Hugh Montgomery (UCH and Whittington Trust) for his encouragement and facilitation. The study was initiated in response to discussions at the National Emergency Critical Care Committee, where Peter McCulloch represents the Chartered Institute of Ergonomics and Human Factors.

